# Individual Differences in the Effects of Neighborhood Socioeconomic Deprivation on Economic Decision Making and Psychotic Risk in Children

**DOI:** 10.1101/2023.04.30.23289335

**Authors:** Junghoon Park, Minje Cho, Eunji Lee, Bo-Gyeom Kim, Gakyung Kim, Yoonjung Yoonie Joo, Jiook Cha

## Abstract

Understanding how a child’s social and economic surroundings influence their mental development and potential for psychological disorders is essential for unpacking the origins of mental health issues. This study, using up-to-date machine learning-based causal inference methods, tested the relationships between neighborhood socioeconomic deprivation, delay discounting, and psychotic-like experiences (PLEs) in 2,135 children considering the wide range of covariates. We found that a greater neighborhood deprivation led to steeper future reward discounting and a higher psychosis risk, evident over 1-year and 2-year follow-ups. We also discovered, across children, significant individual differences in the effect of neighborhood adversity on childhood PLEs, particularly hallucinational symptoms. Children particularly vulnerable to PLEs in adverse neighborhoods exhibited steeper future reward discounting, higher cognitive performance polygenic scores, notable neuroanatomical alterations, including reduced volume, surface area, and white matter in limbic regions. Furthermore, these children displayed increased BOLD reactivity within the prefrontal-limbic system during Monetary Incentive Delay tasks across various reward/loss versus neutral conditions. These findings underscore the intricate interaction between the brain’s reward processing mechanisms and external socioeconomic elements in shaping the risk of psychosis in children.

## Introduction

In *Critique of Practical Reason*, Immanuel Kant champions the inherent power of human reason, suggesting that it is an *a priori* capacity independent of external factors, enabling individuals to engage in responsible actions^1^. Nevertheless, a wealth of scientific studies in recent decades stands in opposition to the Enlightenment philosopher’s claims, highlighting the significant impact of environmental factors on the development of personal identity and behavior.

Adverse childhood environments, such as low family income, malnutrition, physical or sexual abuse, and unsafe neighborhoods, are linked to an heightened risk of various mental or physical health issues, including psychosis^2–4^, impoverished cognitive ability^5–7^, anxiety, bipolar disorder, self-harm, depression^3,4,8^, substance abuse, and obesity^9,10^. Furthermore, these environments are associated with negative social outcomes, such as poor academic performance^11,12^, low income, unemployment^13–18^, higher rates of imprisonment, and increased likelihood of teen pregnancy^19^. Additionally, exposure to these adverse conditions in childhood is associated with a propensity for engaging in risky behaviors, including criminal activity^20^, excessive consumption of calorie-dense foods^21^, substance use^22,23^, deficient self-control^24^, and disrupted reward processing^25^.

The intricate relationship between challenging childhood environments, irresponsible behavior, and adverse social and health outcomes raises important questions. We hypothesized that childhood adversity causes impairment in one’s valuation system, leading to negative life outcomes. Children who experienced social adversities such as poverty show steeper discounting of future rewards in adulthood and have greater risk of psychosis^2,3,26–28^.

Lower socioeconomic status positively correlates with functional brain activity concordance and grey matter volume within reward-related areas (i.e., ventral striatum, putamen, caudate nucleus, orbital frontal cortex) and negatively with executive-related areas (i.e., frontal, medial frontal cortex)^29^. A recent study reported that neuroanatomical features including total cortical volume, surface area, and thickness mediates the association of environmental risk factors and psychotic-like experiences (PLEs) in children^3^.

In addition, individuals with steeper discounting of future rewards (i.e., value present rewards much higher than future rewards) were inclined to save less, invest less in education, more likely to engage in criminal behavior, exhibit lower academic performance, and have less economic wealth^30–33^. This impairment in the intertemporal valuation system is associated with psychiatric disorders, including psychosis, attention deficit/hyperactivity disorder (ADHD), and addiction^34,35^. Psychosis, in particular, has been consistently linked to steep delay discounting^36–38^. Individuals with psychosis may manifest as a skewed neural response to non-relevant rewards, possibly due to increased tonic dopamine^34,39–41^. Blunted dopaminergic projections from the ventral tegmental area to the mesocorticolimbic regions disrupt reward anticipation and perception^39,40^, potentially causing delusions or hallucinations.

In the present study, our primary objective was to investigate the impact of neighborhood socioeconomic deprivation on adolescents’ delay discounting and PLEs. Delay discounting, which is evidenced by the extent to which individuals’ discount future rewards, pertains to their intertemporal decision-making and impulsive behavior. Exposure to adversities at the neighborhood level during childhood has been shown to negatively influence neurocognitive development^7,42,43^, subsequently resulting in psychiatric disorders^2,3,28^ and unfavorable social outcomes, such as decreased income, reduced probability of college attendance, and limited employment opportunities^17^. This phenomenon is particularly pronounced in societies where discrimination based on family income or race/ethnicity restricts underprivileged families from selecting neighborhoods that present greater opportunities for upward social mobility, as observed in the United States^17^.

It is crucial to note that PLEs, frequently reported in children, are considered as a clinically significant risk indicator for psychosis and general psychopathology^44,45^. Around 17% of 9-12 years old children report PLEs^46^, and individuals with PLEs at age 11 had greater risk of developing psychotic disorders in adulthood^47,48^. Prior studies revealed that PLEs are correlated to heightened vulnerability to other psychopathologies including suicidal behavior^2^, mood, anxiety, and substance disorders^44,46^, and exhibit the strongest association with environmental risk factors in comparison to other internalizing/externalizing symptoms during early adolescence^3^. The present study endeavors to explore the potential causal mechanisms underlying these associations.

Our second aim was to test whether the causal effects of neighborhood deprivation on children’s PLEs are heterogeneous based on individual’s delay discounting and its genetic, neural correlates. The heterogeneous nature of psychopathology has long posed significant challenges for clinical diagnosis and treatment^49,50^. Given that the genetic and neural correlates of delay discounting substantially overlap with those of psychosis^40,41,51,52^, the shared biological foundations between reward valuation and psychosis may result in heterogeneous effects of environmental exposure on an individual’s PLEs. By investigating these potential variations, this study seeks to enhance the understanding of the complex interplay between environmental factors and individual predispositions in the development of psychopathology.

Identifying individual differences of treatment/exposure is crucial for the development of personalized health care. Delivering optimal health care for each patient necessitates the recognition of genetic markers, neurodevelopmental characteristics, and sociodemographic features associated with individual variations in treatment effects^53,54^. However, previous studies employing traditional methods of testing the individual differences in treatment effects have often been unsuccessful in discerning the intricate interplay between genetic and environmental factors^55,56^. Linear models with interaction terms of features selected a priori by the researcher may not fully reflect the complex and elusive gene-environment interplay, particularly in genetic and neuroscience research where the input features are usually high dimensional.

Using an up-to-date causal machine learning approach^57,58^, we assessed the effects of neighborhood socioeconomic adversity on delay discounting and PLEs, and the potential individual differences within those effects. We leveraged multimodal magnetic resonance imaging (MRI) data from 11,876 preadolescent children aged 9 to 12 years old (the Adolescent Brain Cognitive Development (ABCD) Study). Integration of innovative analytical techniques and a large sample with diverse genetic and environmental backgrounds permits us to test the complex interactions between genetic and environmental factors, ultimately contributing to the development of more effective personalized health care strategies.

## Results

The demographic characteristics of the final sample (N=2,135) are presented in **Table 1**. Within the sample, 46.14% were female, 76.63% of participants had married parents, the mean family income was $70,245, and 65.57% identified their race/ethnicity as white. To ensure the representativeness of the final sample, a supplementary table comparing the sample’s demographic characteristics with those of the general United States population is provided in the **Supplementary Information** (**Supplementary Table 1**). This comparison serves to reinforce the validity and generalizability of the study’s findings.

**Table 1.**
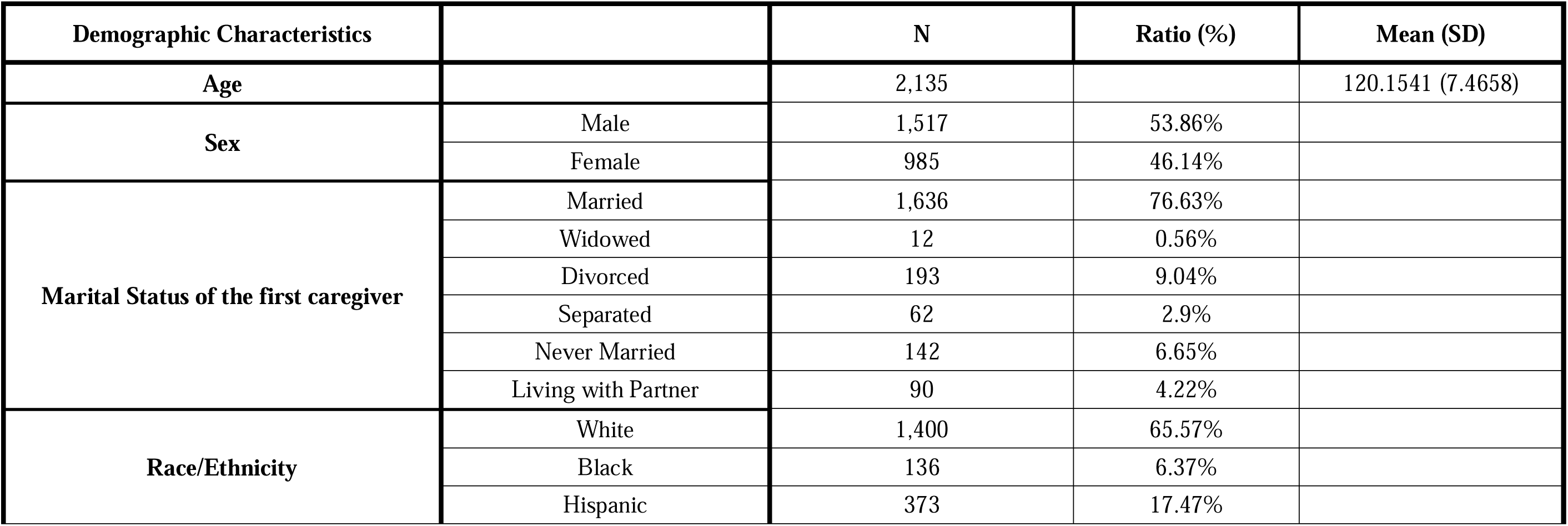
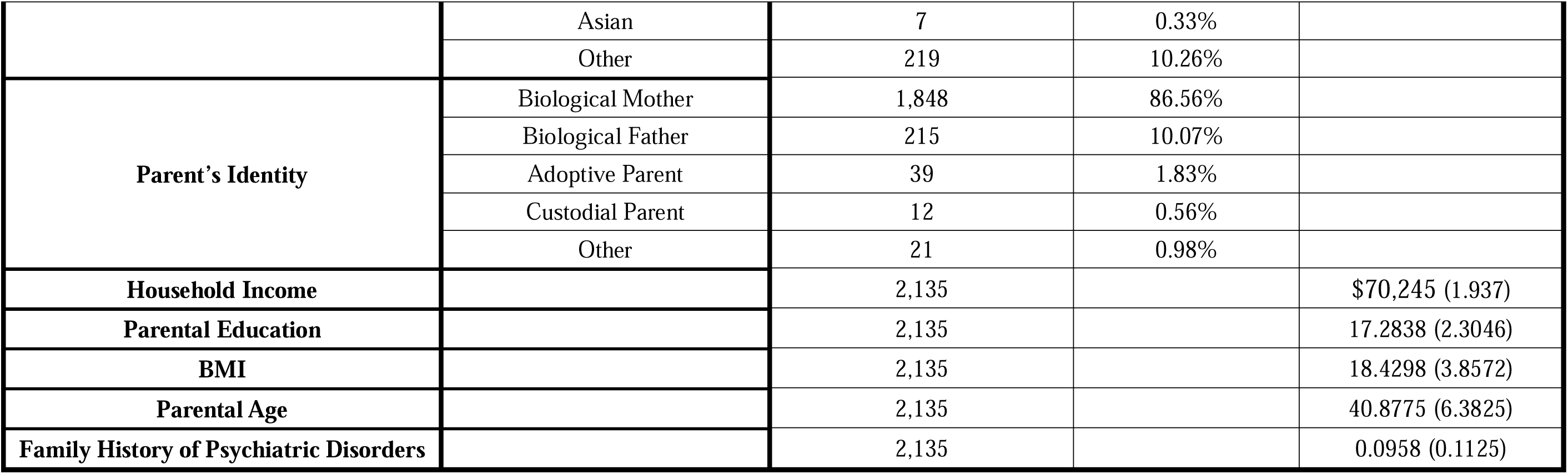
Socioeconomic/demographic characteristics of the participants. *Age* is rounded to chronological month. *Race/Ethnicity* denote child’s self-reported racial / ethnic identity. *Household Income* is assessed as the total combined family income for the past 12 months. *Parental Education* is measured as the highest grade or level of school completed or highest degree received. *Family History of Psychiatric Disorders* represents the proportion of first-degree relatives who experienced mental illness.

In this non-randomized observational study, we addressed potential confounding factors, such as genetic, environmental variables, and their unobserved common causes, which can lead to biased estimations in studies like the ABCD study^59^. We used instrumental variable (IV) random forests (henceforth IV Forest)^57,58^—a random forest-based IV regression^60^—to adjust for unobserved confounding bias in identifying the causal effects of neighborhood socioeconomic adversity (measured with *Area Deprivation Index*, henceforth ADI) on delay discounting and PLEs. The IV Forest method enabled us to derive nonparametric, doubly robust estimates of the average (group-level) and heterogeneous (individual-level) treatment effects of ADI on these outcomes. Notably, this method is particularly useful for analyzing the complex, nonlinear interactions between genetic and environmental factors and their effects on neurocognitive development and psychosis risk, even within the confines of observational data^57^.

**Fig. 1** presents the analytical framework of our study, examining the effects of neighborhood socioeconomic adversity on children’s decision-making and mental health. ADI, recorded in the baseline year, serves as an indicator of this socioeconomic adversity. We assessed the impact of ADI on children’s intertemporal decision-making through delay discounting at a 1-year follow-up. PLEs, encompassing distress, delusional, and hallucinational symptoms, were evaluated at both 1-year and 2-year follow-ups. Our analysis spans multiple follow-up periods and PLE indicators to investigate the sustained influence of ADI over time and to explore differential effects on various PLE symptoms, particularly delusional versus hallucinational.

**Fig. 1.**
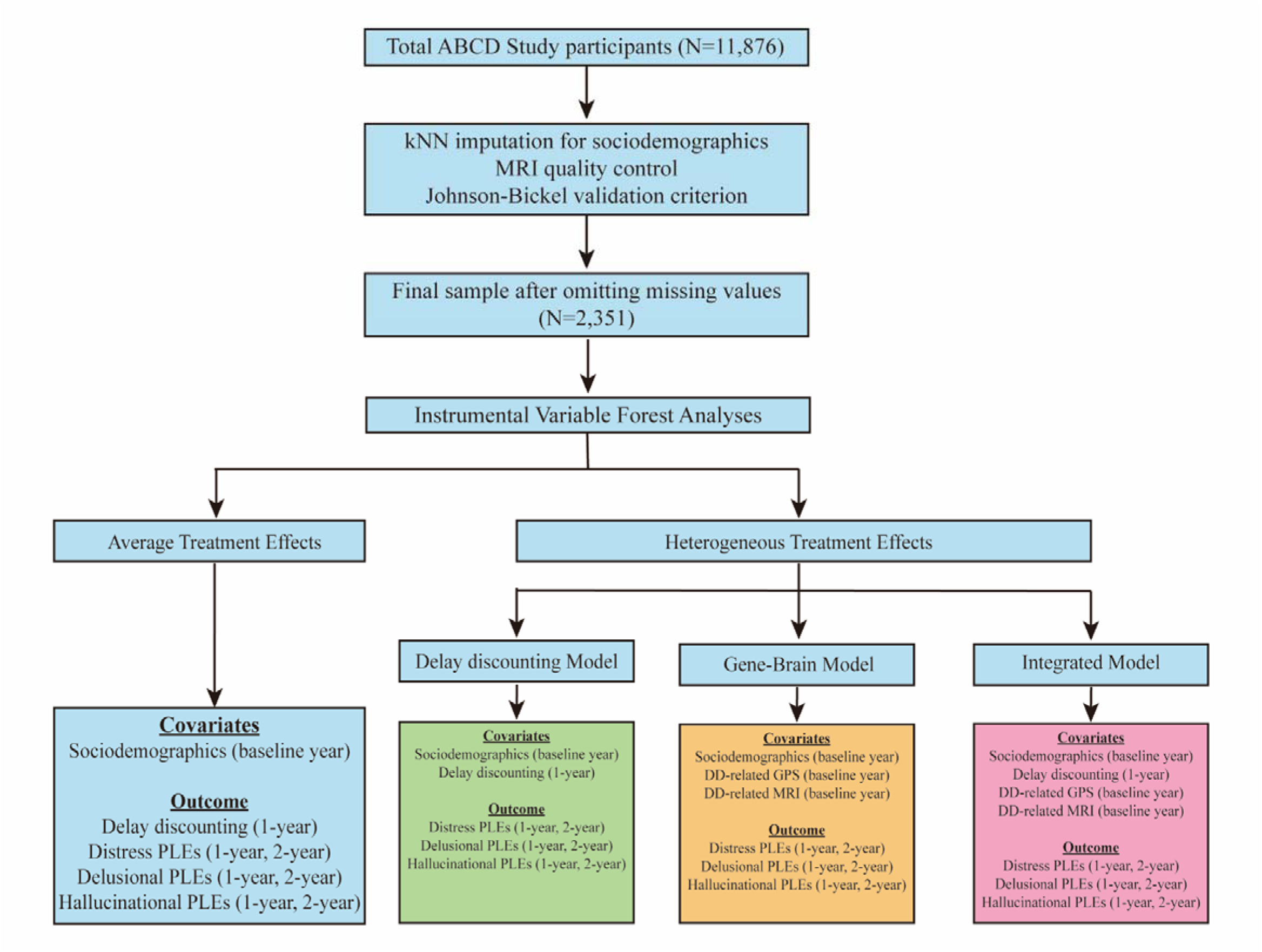
Study flow diagram. This figure illustrates the participant selection and data processing in our study. We initially included 11,876 participants aged 9-12 years from the Adolescent Brain Cognitive Development (ABCD) Study, utilizing the release 4.0 dataset which encompasses baseline, 1-year follow-up, and 2-year follow-up observations. Sociodemographic features underwent kNN imputation. Subsequently, we excluded observations not meeting the ABCD Study’s MRI quality control standards and those failing the Johnson-Bickel validation criterion for delay discounting. This resulted in a final sample size of N=2,351. Using this cohort, our study first investigated the average treatment effects of neighborhood socioeconomic deprivation on children’s intertemporal valuation of rewards and psychotic risks. We then explored the individual differences of these effects in relation to children’s delay discounting behaviors and associated genetic and neural factors.

### Average Treatment Effects of Neighborhood Socioeconomic Adversity on Delay Discounting and PLEs

IV Forest analyses revealed that a higher ADI has significant (causal) associations with a lower delay discounting (β= -1.73, p-FDR= 0.048) and a higher PLEs (distress score 1-year follow-up: β= 1.872, p-FDR= 0.048; distress score 2-year follow-up: β= 1.504, p-FDR= 0.039; delusional score 1-year follow-up: β= 5.97, p-FDR= 0.048; delusional score 2-year follow-up: β= 4.022, p-FDR=0.048; hallucinational score 1-year follow-up: β= 3.761, p-FDR= 0.048; hallucinational score 2-year follow-up: β= 4.786, p-FDR=0.039) (**Table 2**).

**Table 2.** Causal effects of neighborhood socioeconomic adversity on intertemporal valuation and psychotic risk. Average treatment effects of ADI on delay discounting and PLEs in the IV Forest models are shown. All p-values were corrected for multiple comparison using false discovery rate.

|  | IV Forests: Average Treatment Effects |  |  |  |  |
| --- | --- | --- | --- | --- | --- |
|  | Estimates | Std. Error | 95% Lower CI | 95% Upper CI | P-FDR |
| Delay Discounting<br>(1-year follow-up) | -1.730 | 0.748 | -3.195 | -0.265 | 0.048 |
| Distress Score PLEs<br>(1-year follow-up) | 1.872 | 0.612 | 0.673 | 3.071 | 0.048 |
| Distress Score PLEs<br>(2-year follow-up) | 1.504 | 0.592 | 0.345 | 2.664 | 0.039 |
| Delusional Score PLEs<br>(1-year follow-up) | 5.970 | 2.911 | 0.264 | 11.676 | 0.048 |
| Delusional Score PLEs<br>(2-year follow-up) | 4.022 | 1.987 | 0.127 | 7.917 | 0.048 |
| Hallucinational Score PLEs<br>(1-year follow-up) | 3.761 | 1.902 | 0.033 | 7.489 | 0.048 |
| Hallucinational Score PLEs<br>(2-year follow-up) | 4.786 | 1.861 | 1.139 | 8.433 | 0.039 |

Supplementary analyses were conducted using a conventional linear IV regression^60^ and an alternative causal machine learning method (i.e., *Double ML*^61,62^) to validate the results. The conventional IV regression also showed that ADI has negative influence on childhood delay discounting (β= -0.468, p-FDR= 0.03) and positive PLEs (distress score 1-year follow-up: β= 0.609, p-FDR= 0.011; distress score 2-year follow-up: β= 0.78, p-FDR= 0.003; delusional score 1-year follow-up: β= 0.486, p-FDR= 0.028; delusional score 2-year follow-up: β= 0.578, p-FDR= 0.013; hallucinational score 1-year follow-up: β= 0.604, p-FDR= 0.011; hallucinational score 2-year follow-up: β= 0.827, p-FDR= 0.003). The partial-linear IV model of the Double ML algorithm showed significant effects of ADI on children’s delay discounting (β= -0.429, p-FDR= 0.044), distress score PLEs (1-year follow-up: β= 0.495, p-FDR= 0.023; 2-year follow-up: β= 0.609, p-FDR= 0.005), hallucinational score PLEs (1-year follow-up: β= 0.498, p-FDR= 0.018; 2-year follow-up: β= 0.683, p-FDR= 0.002), and 2-year follow-up delusional score PLEs (β= 0.417, p-FDR= 0.044). The negative effects of ADI on 1-year follow-up delusional score PLEs were marginally significant (β= 0.393, p-FDR= 0.051). These results of the conventional linear IV regression and Double ML partial-linear IV regression confirm the findings obtained from the IV Forest (**Supplementary Table 3**), further supporting the primary analyses and conclusions drawn from the study.

### Heterogeneous Treatment Effects of Neighborhood Socioeconomic Adversity on PLEs, conditioned on the Genetic and Neural Correlates of Delay Discounting

Next, we tested whether the impact of ADI was heterogeneous across children, and, if so, whether the heterogeneity is linked to individual’s neurodevelopmental characteristics and the relevant genetic factors—assessed with genome-wide polygenic scores (GPS) and structural MRI and monetary incentive delay (MID) task fMRI data—correlated to intertemporal valuation. To identify the best subset of genetic and neural correlates of delay discounting, we first selected GPS and MRI brain regions of interest (ROIs) specifically related to delay discounting. To analyze the nonparametric correlations of multiple input variables, we used a random forest-based feature selection *Boruta* algorithm^63^. Its robustness and effectiveness in selecting relevant features in high dimensional, intercorrelated biomedical data (e.g., MRI) has been validated^63^ and consistently applied in genetics and neuroscience research^64–66^. The variables significantly correlated with delay discounting (p-Bonferroni<0.05) were GPS of cognitive performance, IQ, and education attainment; morphometric features (e.g., surface area, volume) in the limbic system (temporal pole, parahippocampal gyrus, caudate nucleus, rostral anterior cingulate, isthmus cingulate), inferior frontal gyrus (pars opercularis), and fusiform gyrus; mean beta activations of rewards/losses versus neutral feedback in the subcortical areas (thalamus proper, ventral diencephalon), precentral gyrus, supramarginal gyrus, temporal lobe (transverse temporal gyrus, superior temporal gyrus), and insula (**Supplementary Table 4**).

We then assessed the heterogeneous treatment effects of ADI on PLEs using three distinct IV Forest models: (1) the Delay Discounting model, incorporating sociodemographic features and delay discounting; (2) the Gene-Brain model, which included sociodemographic features and genetic and neural correlates of delay discounting (i.e., GPS and brain ROIs identified using the Boruta algorithm); and (3) the Integrated model which combined all the variables from the previous two models. All three models satisfied the overlap assumption (i.e., the estimated propensity scores are not close to one or zero), which is crucial for the validity of the estimated heterogeneous treatment effects (**Supplementary Fig. 1**). In line with prior studies^67,68^, we obtained conditional average treatment effects, divided subjects into deciles (Q1: most vulnerable; Q10: most resilient) based on the conditional average treatment effects, and conducted three hypothesis tests^69^ on each model to determine the most effective model for capturing the individual differences (heterogeneity) in the effects of ADI on PLEs: monotonicity, alternative hypothesis, and ANOVA.

Among the three models, only the Integrated model successfully demonstrated significant individual differences in the ADI effects on PLEs. This was evident in the impact of ADI on 1-year follow-up distress score PLEs (monotonicity test: p-FDR=0.011; alternative hypothesis test: p=0.002; ANOVA test: p<0.001) and 1-year follow-up hallucinational score PLEs (monotonicity test: p-FDR=0.038; alternative hypothesis test: p=0.004; ANOVA test: p<0.001), as presented in **Fig. 2 and Table 3**. In contrast, the Delay Discounting model and Gene-Brain model failed to pass the heterogeneity tests (monotonicity test: p-FDR≥0.05; alternative hypothesis and ANOVA test: p≥0.05).

**Fig. 2.**
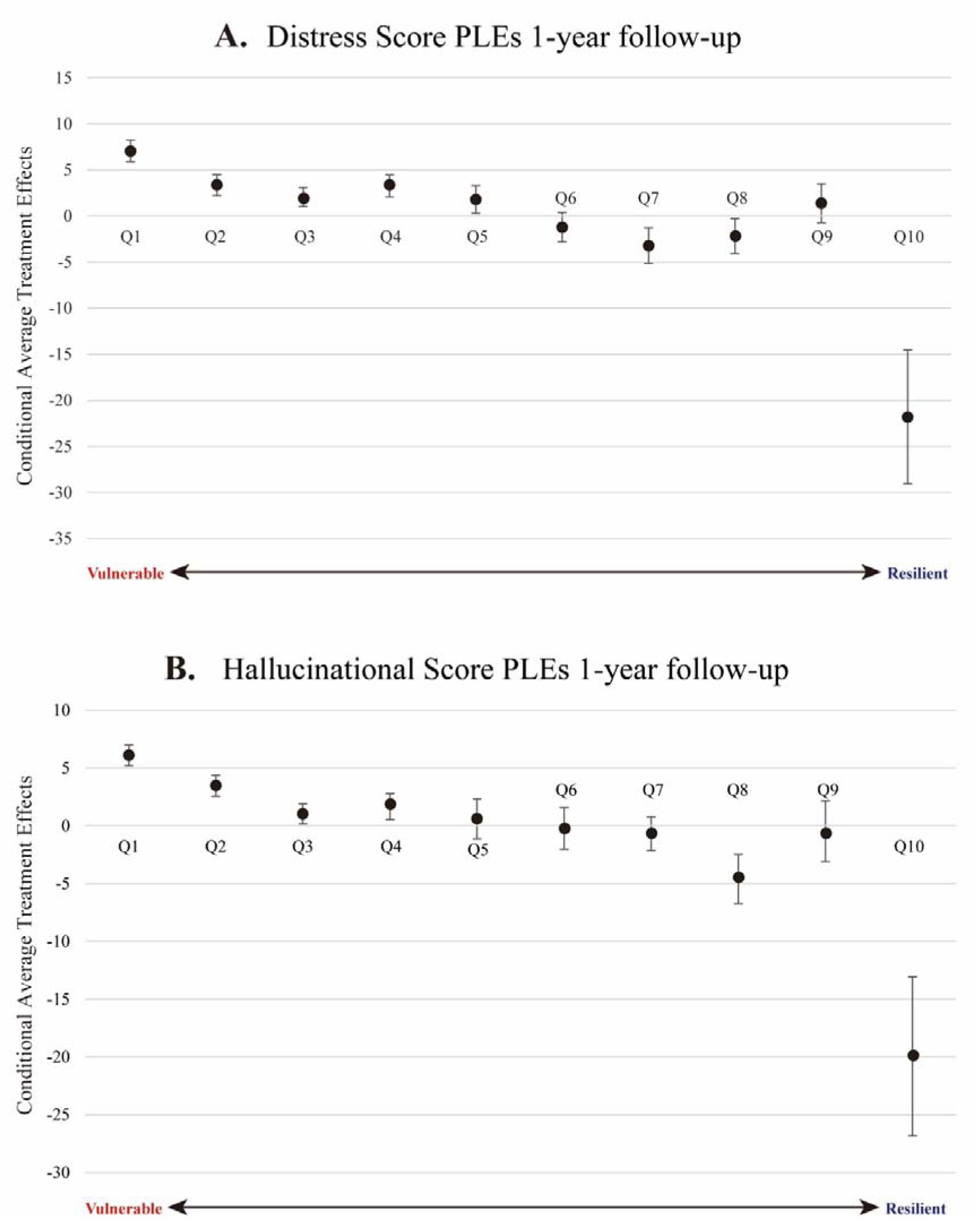
Delineation of heterogeneous treatment effects by vulnerable/resilient subgroups. Heterogeneity in the average treatment effects of the ADI on PLEs are shown as a bar plot, specifically focusing on 1-year follow-up distress score PLEs (**A**) and hallucinational score PLEs (**B**). These effects are plotted across ten deciles, which are organized based on the relative vulnerability or resilience of the participants, with Q1 denoting the most vulnerable and Q10 indicating the most resilient. Point estimates of the conditional average treatment effect of each decile were derived via a doubly-robust estimation method within the IV Forest algorithm. 95% confidence intervals are depicted using error bars.

**Table 3.** Evaluation of individual differences in the causal effects of neighborhood socioeconomic adversity on children’s psychotic risk. We employed three distinct tests to evaluate the heterogeneous treatment effects of ADI on PLEs: monotonicity test, alternative hypothesis test, and ANOVA test. These tests were applied to three developed IV Forest models: Delay Discounting model, Gene-Brain model, and Integrated model. All p-values were corrected for multiple comparison using false discovery rate. A star (*) denotes overall significance, indicating that a model passed all three heterogeneity tests.

|  |  | Monotonicity | Alternative Hypothesis |  |  |  | ANOVA |  | Overall Significance |
| --- | --- | --- | --- | --- | --- | --- | --- | --- | --- |
|  |  | p-FDR | Estimate | Std. Error | t-value | p-FDR | F-value | p-FDR |  |
| **Delay Discounting Model** | Distress Score PLEs (1-year follow-up) | 0.037 | -22.261 | 40.164 | -0.554 | 0.799 | 1.114 | 0.48 |  |
|  | Distress Score PLEs (2-year follow-up) | 0.094 | -54.769 | 142.528 | -0.384 | 0.799 | 1.081 | 0.48 |  |
|  | Delusional Score PLEs (1-year follow-up) | 0.019 | 55.047 | 188.400 | 0.292 | 0.799 | 0.965 | 0.495 |  |
|  | Delusional Score PLEs (2-year follow-up) | 0.303 | 23.289 | 91.550 | 0.254 | 0.799 | 0.852 | 0.568 |  |
|  | Hallucinational Score PLEs (1-year follow-up) | 0.362 | -86.756 | 241.597 | -0.359 | 0.799 | 0.995 | 0.495 |  |
|  | Hallucinational Score PLEs (2-year follow-up) | 0.098 | -119.595 | 341.060 | -0.351 | 0.799 | 1.010 | 0.495 |  |
| **Gene-Brain**<br>**Model** | Distress Score PLEs<br>(1-year follow-up) | 0.058 | -9.522 | 4.244 | -2.244 | 0.05625 | 2.289 | 0.027 |  |
|  | Distress Score PLEs<br>(2-year follow-up) | 0.099 | -9.202 | 7.016 | -1.312 | 0.285 | 1.608 | 0.161 |  |
|  | Delusional Score PLEs<br>(1-year follow-up) | 0.086 | -7.463 | 3.262 | -2.288 | 0.05625 | 2.631 | 0.01 |  |
|  | Delusional Score PLEs<br>(2-year follow-up) | 0.043 | -9.512 | 4.886 | -1.947 | 0.0936 | 3.493 | 0.001 |  |
|  | Hallucinational Score PLEs<br>(1-year follow-up) | 0.022 | -8.917 | 4.123 | -2.163 | 0.062 | 1.878 | 0.083 |  |
|  | Hallucinational Score PLEs<br>(2-year follow-up) | 0.146 | -9.382 | 5.681 | -1.651 | 0.162 | 2.695 | 0.009 |  |
| **Integrated**<br>**Model** | Distress Score PLEs<br>(1-year follow-up) | 0.011 | -8.922 | 2.922 | -3.053 | 0.0252 | 8.388 | <0.001 | * |
|  | Distress Score PLEs<br>(2-year follow-up) | 0.074 | -9.343 | 3.469 | -2.693 | 0.0252 | 7.361 | <0.001 |  |
|  | Delusional Score PLEs<br>(1-year follow-up) | 0.172 | -6.946 | 2.460 | -2.824 | 0.0252 | 6.816 | <0.001 |  |
|  | Delusional Score PLEs<br>(2-year follow-up) | 0.136 | -6.854 | 2.617 | -2.619 | 0.027 | 7.525 | <0.001 |  |
|  | Hallucinational Score PLEs<br>(1-year follow-up) | 0.038 | -8.246 | 2.834 | -2.910 | 0.0252 | 7.182 | <0.001 | * |
|  | Hallucinational Score PLEs<br>(2-year follow-up) | 0.089 | -8.355 | 3.070 | -2.721 | 0.0252 | 9.484 | <0.001 |  |

To elucidate the role of specific genetic and neural correlates within the heterogeneous effects of ADI on PLEs, we obtained Shapley additive explanation (SHAP) scores^70^. SHAP scores provide insights into how each variable contributes positively or negatively to the differential causal effects of ADI on 1-year follow-up observations of distress score and hallucinational score PLEs. These scores help differentiate the roles of these factors across subgroups, ranging from low to high conditional average treatment effects, thereby providing a nuanced understanding of how ADI influences PLEs through various genetic and neural pathways.

In both distress score and hallucinational score PLEs, children who showed higher levels of ADI’s adverse effects on PLEs exhibited distinct neuroanatomical and functional brain patterns, particularly in the limbic system. These patterns included reduced neuroanatomical features such as smaller white matter and surface area in the right temporal pole, reduced area and volume in the right parahippocampal region, decreased left white surface area, smaller area in the right isthmus cingulate, reduced intracranial volume, smaller caudate nucleus volume, and lower total grey matter volume. Functionally, greater activation during MID tasks was observed in several areas including the posterior cingulate, right ventral diencephalon, right insula, left thalamus proper, and left precentral gyrus. Additionally, children more adversely affected by ADI, as indicated by higher conditional average treatment effects on PLEs, exhibited larger right fusiform volume, decreased activation in the left superior temporal gyrus, younger parental age, and lower BMI (**Fig. 3**).

**Fig. 3.**
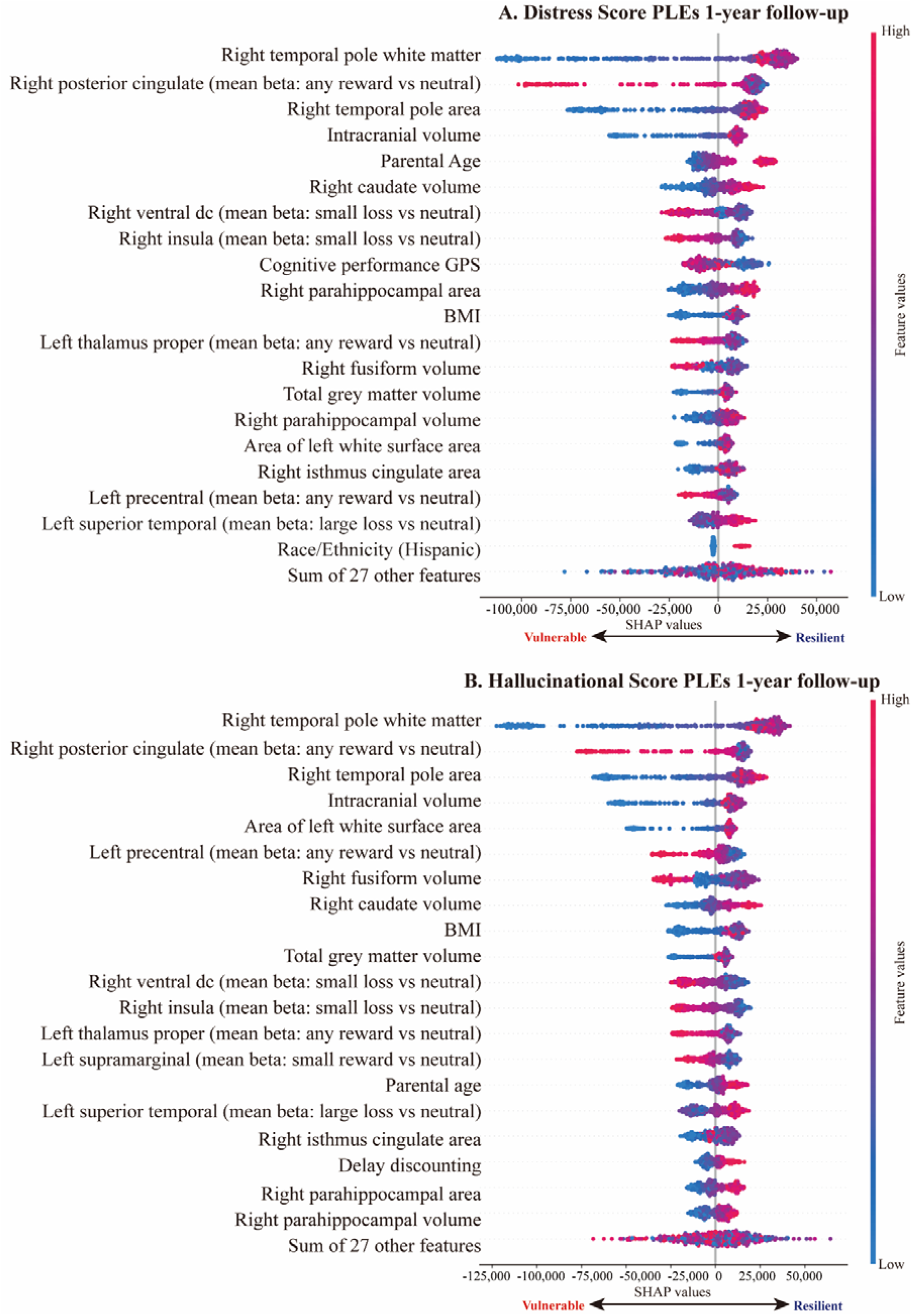
Beeswarm summary plots of Shapley additive explanation (SHAP) values for Integrated model. Contributions of the top 20 variables of highest importance in the Integrated model for the heterogeneous treatment effects of neighborhood socioeconomic deprivation on 1-year follow-up distress score PLEs (**A**) and 1-year follow-up hallucinational PLEs (**B**) are shown. Variables are ordered by their relative importance in the model. Negative SHAP values indicate greater vulnerability (lower resilience) to the effects of ADI on PLEs; Positive values indicate lower vulnerability (greater resilience). Contrasts of average beta activations of the given brain ROIs during MID tasks are shown in parenthesis. GPS: genome-wide polygenic scores; Ventral dc: ventral diencephalon.

The analysis also revealed that higher conditional average treatment effects on distress score PLEs was associated with higher cognitive performance GPS and a lower likelihood of being Hispanic. In contrast, for hallucinational score PLEs, greater importance was attributed to increased activation in the left supramarginal gyrus during MID tasks and more pronounced discounting of future rewards. These nuanced associations are depicted in **Fig. 3**.

Lastly, we conducted a supplementary analysis to test whether the effects of delay discounting between the causal impact of ADI on PLEs are captured with a conventional linear mediation model^71^. This linear IV mediation model showed no significant mediation effects of delay discounting (β= -6.929E-6 [95% CI, -0.012∼0.026] ∼ 4.582E-6 [95% CI, - 0.009∼0.03]) (**Supplementary Table 5**).

## Discussion

In this study, we examined how neighborhood socioeconomic deprivation impacts children’s intertemporal choice behavior (delay discounting) and psychotic risk, considering the multifaceted effects of neighborhood adversity and its underlying biological, environmental, and behavioral drivers. Our findings can be distilled into two main points. Firstly, there was a notable link of living in socioeconomically disadvantaged neighborhoods to the propensity for children to prefer immediate rewards over larger, delayed ones—a behavior known as steep delay discounting (indicative of lower impulse control) and to a higher rate of PLEs. This association was significant even after adjusting for a range of confounding factors, both observed (e.g., familial socioeconomic status) and unobserved. Secondly, the influence of disadvantaged neighborhood environments on PLEs was found to be heterogeneous. This individual variability is influenced not just by delay discounting, but also by a confluence of factors including genetic predisposition for cognitive intelligence, and brain morphometry and functioning (task activation). Causal machine learning models utilized in our study have identified a spectrum of conditions that either exacerbate vulnerability or contribute to resilience, accounting for the diverse effects of neighborhood environments on children’s risk of developing psychosis.

Our findings hold implications for social science. Using causal machine learning models, such as IV Forest and Double ML, we provide consistent and clear results that residential adversity during childhood leads to steeper discounting of future rewards. This finding challenges the longstanding economic theory that an individual’s rate of discounting future rewards (time preference) is an exogenous parameter of intertemporal choice, established a priori, and impervious to external influences^32^. Since the introduction of the discounted utility model^72^ by Paul Samuelson in 1937, there has been limited exploration into whether environmental factors affect the development of an individual’s parameter^32,73^. Although recent studies has hinted at the impact of socioeconomic status^33,74–77^ and cultural norms^73,78^ on one’s intertemporal decision making, however, the causal mechanisms have remained elusive. Our study builds on these inquiries, offering concrete evidence that the development of an individual’s time preference is subject to environmental influences, and thereby opening new avenues for understanding the dynamics of intertemporal decision-making.

We address this knowledge gap by identifying the potential causal influence of neighborhood environment on intertemporal choice, leveraging longitudinal observations of preadolescent children aged 9-12 years, a critical period for neurocognitive development. Given that an individual’s intertemporal valuation of rewards contributes to economic and health disparities between individuals^30,34,52^, early socioeconomic deprivation may result in a behavioral poverty trap^33^. In such scenarios, individuals raised in impoverished environments are prone to shortsighted behavior, exacerbating the challenge of escaping poverty. A plausible mechanism for this phenomenon is the effects of glucocorticoid on brain’s reward system. Prior studies indicate that adverse social environments induce chronic stress to children, elevating glucocorticoid hormones like cortisol^79–83^. In particular, neighborhood socioeconomic deprivation has a more pronounced association with cortisol increases in children compared to any other social environmental factors^84^. Long-term chronic stress from growing up in disadvantaged neighborhoods could result in epigenetic modifications affecting the mesocorticolimbic dopaminergic system, thereby altering the reward system^82,83^. This alteration may lead to a heightened preference for immediate rewards and impulsive behaviors, such as unhealthy eating and substance abuse^81–83,85–89^, further entrenching the cycle of socioeconomic disadvantage.

Our second findings extend this understanding by linking the heterogeneous effects of ADI on children’s PLEs with the intricate relationship between childhood social adversity and the reward system. Our findings suggest that these differential effects of neighborhood socioeconomic adversity are modulated by genetic predispositions and neurodevelopmental traits associated with delay discounting. Children who experience residential deprivation and are at a higher psychotic risk demonstrate several distinct characteristics, including lower BMI, younger parental age, and altered brain structures and functions associated with delay discounting. Notably, these children showed reduced volume or white matter in specific brain regions (right temporal pole, right parahippocampal gyrus, right caudate nucleus, right isthmus cingulate), along with a smaller intracranial and total grey matter volume. Functionally, these children showed greater activation during MID tasks in regions including the right posterior cingulate, right ventral diencephalon, right insula, left precentral gyrus, left thalamus proper, and left superior temporal. This is particularly pronounced in children with a greater propensity for hallucinatory symptoms, who also show increased activity in the left supramarginal gyrus.

It appears that variations in structural and functional aspects of the limbic system (the posterior cingulate, ventral diencephalon, insula, temporal pole, parahippocampal gyrus, and isthmus cingulate) play a crucial role in how socioeconomic hardship affects PLEs. This individual variability may be linked to individual differences in the glucocorticoid and reward system. The interaction between our genes and neurons, in response to chronic stress from poor socioeconomic conditions, may determine the differing impacts of such adversity on PLEs.

Although direct testing of this association within the ABCD Study samples was not feasible due to lack of relevant data, extensive animal and human corroborate our hypotheses. These studies suggest that maladaptive valuation of intertemporal rewards, namely the excessive discounting of future rewards, is linked to dysfunction of the prefrontal-limbic system, associated with psychopathologies such as psychosis in adolescents and adults^34,35,38–41,90^. Animal models have demonstrated that adverse social environments trigger chronic dysregulation of glucocorticoid signaling in the hypothalamic-pituitary-adrenal axis and the dopaminergic mesocortical circuit^83^, through epigenetic control^82,83^. This dysregulation disrupts the adolescent reward circuit. In humans, childhood exposure to social adversity leads to changes in the hypothalamic-pituitary-adrenal axis and contributes to psychosis through abnormal neurodevelopment of the limbic regions, the temporal pole, cingulate cortices, parahippocampal gyrus, and caudate nucleus^91–94^. Young adults with a history of childhood social deprivation often show impaired reward processing, particularly in the cingulate and mesostriatal dopaminergic system^25,94–96^.

The age of our study’s participants, 9-12 years old, is a critical period for development of the prefrontal-limbic system^94,97,98^. Children with psychotic disorders often exhibit greater reductions in grey matter and lower synaptic density compared to their healthy peers^99,100^. These neurodevelopmental alterations are associated with increased neuronal excitation, reduced inhibitory neural activities, and the resultant impulsive behaviors^101^. In line with our findings, previous research has shown a correlation between higher psychotic risk and neuroanatomical alterations in the right temporal fusiform, right temporal pole, and right parahippocampal gyrus^93^, as well as greater neural activations in limbic regions such as the insula and cingulate cortices during reward outcomes in MID task^102^. Overall, our findings on the heterogeneous effects of neighborhood deprivation contribute to the growing body of literature showing the role of glucocorticoid and reward systems in modulating the adverse effects of environmental deprivation on psychosis^92,94,96,103,104^.

In our study, we discovered that children, when exposed to deprived neighborhoods and already facing the challenges of residential disadvantage, were more likely to experience PLEs. Surprisingly, these children also showed a higher GPS for cognitive performance. At first glance, this finding seems to contradict prior research, which has consistently identified a negative relationship between PLEs and cognitive performance^28,105^.

To understand this complex relationship, we turned to the bioecological model and the Scarr-Rowe hypothesis on gene-environment interactions^106–108^. This theory proposes that the impact of genetic factors is lessened in unfavorable environments. An easy way to visualize this is by comparing it to plant growth: in poor soil, a plant can’t get the nutrients it needs, which limits its growth despite its genetic potential to grow tall^109^. But, when these children face residential disadvantages, this protective gene-psychosis link weakens. Their genetic resilience decreases, making them more vulnerable to the negative impacts of such disadvantages on PLEs. Essentially, those with higher cognitive ability GPS lose more of their potential genetic protection, making them more susceptible to the adverse effects of their environment on PLEs.

Consistent with our findings, recent large-scale studies have demonstrated that the impact of genetics on brain structure, cognitive functions, and mental health disorders becomes less significant in harmful environments (e.g., abuse)^110,111^. Conversely, in more supportive and enriched settings, like those associated with higher socioeconomic status, genetic influences are more noticeable (e.g., high socioeconomic status)^107,112,113^. Together with these findings, our study contributes to a deeper understanding of how genetic and environmental factors interact to influence the development of psychopathology in children.

In this study, we utilized innovative causal machine learning techniques to test the negative impacts of neighborhood deprivation on childhood psychopathology. Specifically, we employed the IV Forest method that allows us to discern how residential deprivation influences children’s risk of psychosis in a manner dependent on a variety of genetic risk factors (e.g., GPS of cognitive performance, educational attainment, and IQ^105,114,115^) and environmental risk factors (e.g., family income^3,116^), as identified in existing literature. Our findings were adjusted to account for potential biases from both observed and unobserved variables.

The machine learning algorithm we used was adept at modelling the complex interplay gene-environment interactions. Among the three IV Forest models we tested (i.e., Delay Discounting, Gene-Brain, Integrated), only the Integrated model—which included delay discounting, sociodemographic characteristics, and genetic and neural correlates of delay discounting—identified the significant heterogeneous effects of ADI on children’s PLEs. This suggests that the intricate interactions among environmental, genetic, neural factors, and delay discounting play a crucial role in how socioeconomic adversity impacts psychotic risk.

In contrast, traditional linear mediation analysis, which relies on predefined interaction terms in a deductive statistical framework, failed to identify any significant mediation effect of delay discounting between neighborhood deprivation and PLEs. This underscores the effectiveness of our advanced causal machine learning approach over conventional methods in detecting the subtle effects of various interacting factors on childhood psychopathology.

The IV Forest model, unlike traditional methods, allows for data-driven feature selection and stratification of heterogeneous treatment effects^57,58^. It inductively identifies nonlinear and complex patterns of these effects, which are not predetermined by researchers, offering a more nuanced understanding of the data. Prior studies relying on the deductive approach often suffer from low statistical power and bias^59,117^, inadequately reflecting the complexity of gene-environment interactions^55,56^. Consequently, we believe that causal modeling approaches that assess heterogeneous treatment effects hold significant potential as powerful tools for advancing precision science in psychology and medicine. These approaches provide a more dynamic and accurate framework for understanding the multifaceted influences on psychopathology, demonstrating significant promise for future research in these fields.

Several limitations of this study warrant consideration. Firstly, interpretations of our findings as true causality should be approached with caution. Our research is based on the ABCD Study, a non-randomized, observational study, making it prudent to be cautious in interpreting these findings as definitively causal. Secondly, since the majority of participants identified their race/ethnicity as white (63.76%, similar to the US population), the generalizability of our findings to other minor race/ethnicity might remain to be tested. Nonetheless, recent research suggests that temporal discounting measures are consistent across diverse populations worldwide (61 countries, n=13,629)^78^, which may mitigate concerns regarding the representativeness of our findings. Thirdly, the short follow-up periods in our study (1-year and 2-year follow-up) may not adequately capture the long-term neurodevelopmental processes underlying intertemporal valuation and related psychopathology. As the ABCD Study continues to collect more longitudinal observations, longer follow-up periods in future studies could yield deeper insights. Fourthly, despite efforts to ensure representativeness by recruiting from diverse school systems across 21 research sites in the United States, our sample does not fully mirror the entire US population^118^. To address this, we provide a supplementary table (**Supplementary Table 1**) comparing the demographic characteristics of our final sample with the general United States population enhancing the relevance and generalizability of our results. Lastly, future research should examine the heterogeneous effects of additional environmental risk factors, such as parenting behavior^28^ and early life stress^111^, to provide a more comprehensive understanding of the environmental influences on psychopathology.

This study highlights the differential effects of neighborhood disadvantage on intertemporal economic decisions and psychotic risk during early childhood. It underscores the importance of identifying diverse treatment effects by integrating genetic and environmental factors to guide personalized healthcare approaches. Furthermore, we propose that enhancing the childhood environment could contribute to the reduction of economic and health inequality gaps. Economic policies promoting positive intertemporal choice (e.g., increased savings, healthy diet) have predominantly focused on paternalistic welfare policies in adulthood. These policies often assume that an individual’s tendency to discount future rewards is fixed (“exogenous”)^32^. However, our findings suggest that policies or interventions aimed at enhancing the socioeconomic environment during childhood may foster improved intertemporal choice behavior, thereby reducing economic^33^ and health inequality^23,119^. By addressing the root of the problem, this indirect approach may assist individuals in developing the capacity to make more informed choices, ultimately promoting better outcomes.

The insights gleaned from our novel analytical methods revive longstanding philosophical inquiries: do humans possess reason or free will independent of their environment? If our ability to act responsibly is indeed shaped by external circumstances, this challenges the traditional rationale for penalizing criminal and morally objectionable behavior based on the assumption of free will. This inquiry underscores the need for further interdisciplinary research, bridging insights from psychology, sociology, neuroscience, ethics, and law, to explore the nuanced relationship between individual agency and environmental influences. Such research is crucial for understanding how external factors impact decision-making and behavior, thereby informing more nuanced approaches to ethical and legal accountability. It invites a reevaluation of responsibility and justice, suggesting that effective interventions and policies must consider the complex interplay of individual predispositions and environmental conditions in shaping behavior.

## Methods

### Study Participants

The ABCD Study recruited participants from 21 research sites across the nation, utilizing a stratified, probability sampling method to capture the sociodemographic variation of the US population^120^. We used the baseline, first year, and second year follow-up datasets included in ABCD Release 4.0, downloaded on February 10, 2022.

Of the initial 11,876 ABCD samples, we removed participants without genotype data, MRI data, NIH Toolbox Cognitive Battery, delay discounting, residential address, ADI, PLEs. As recommended by the ABCD team^121^, Johnson & Bickel’s two-part validity criterion^122^ was used to exclude subjects with inconsistent responses (i.e., indifferent point for a given delay larger than that of an indifference point for a longer delay). Missing values of covariates were imputed using k-nearest neighbors. The final samples included 2,135 multiethnic children.

### Data

#### Neighborhood Disadvantage

Neighborhood disadvantage was measured with Residential History Derived Scores based on the Census tracts of each respondent’s primary addresses by the ABCD team. Consistent with prior research^3,42^, we chose national percentile scores of the Area Deprivation Index, calculated from the 2011∼2015 American Community Survey 5-year summary. It has 17 sub-scores regarding various socioeconomic factors such as median household income, income disparity, percentage of population aged more than 25 years or more with at least a high school diploma, and percentage of single-parent households with children aged less than 18 years, etc. Higher values of the Area Deprivation Index and poverty and fewer years of residence indicate greater residential disadvantage.

#### Delay Discounting

Delay discounting was measured by the adjusting delay discounting task in the 1-year follow-up ABCD data^121,123^. Each child was asked to make choices between a small immediate hypothetical reward or a larger hypothetical $100 delayed reward at multiple future time points (6h, one day, one week, one month, three months, one year, and five years). By increasing or decreasing the smaller immediate reward depending on the child’s response, the task records the indifference point (i.e., the small immediate amount deemed to have the same subjective value as the $100 delayed reward) at each of the seven delay intervals. Test-retest reliability of this delay discounting measure has been validated^124,125^. Studies show that preadolescent children are capable of comprehending the delay discounting task and show similar patterns of discounting as adults^126^.

To avoid methodological problems regarding mathematical discounting models (hyperbolic vs. exponential) and positively skewed parameters of discounting functions^125,127^, we used the area under the curve, a model-free measure of delay discounting^127^. The area under the curve measure of delay discounting rates (henceforth *discount rates*) ranges from 0 to 1, with lower values indicating steeper discounting and higher impulsivity.

#### Psychotic-Like Experiences

First and second-year follow-up observations of psychotic-like experiences (PLEs) were measured using the Prodromal Questionnaire-Brief Child Version (PQ-BC; child-reported). PQ-BC has a 21-item scale validated for use with a non-clinical population of children aged 9-10 years^128,129^. In line with the previous research^3,114,128,129^, we computed *Total Score* and *Distress Score*, each indicating the number of psychotic-like symptoms and levels of total distress. Total Score is the summary score of 21 questions ranging from 0 to 21, and Distress Score is the weighted sum of responses with the levels of distress, ranging from 0 to 126. Additionally, to test whether the heterogeneous treatment effects of neighborhood adversity differ among psychotic symptoms, Distress Score was divided into two separate scores: *Delusional Score* and *Hallucinational Score*^2,130^. A higher value indicates greater severity of PLEs.

#### Genome-wide Polygenic Scores

Children’s genetic predispositions were assessed with genome-wide polygenic scores (GPS). Summary statistics from genome-wide association studies were used to generate GPS of cognitive intelligence (cognitive performance^131^, education attainment^131^, IQ^132^), psychiatric disorders (major depressive disorder^133^, post-traumatic stress disorder^134^, attention-deficit/hyperactivity disorder^135^, obsessive-compulsive disorder^136^, anxiety^137^, depression^138^, bipolar disorder^139^, autism spectrum disorder^140^, schizophrenia^141^, cross disorder^142^), and health and behavioral traits (BMI^143^, neuroticism^144^, worrying^144^, risk tolerance^145^, automobile speeding propensity^145^, eating disorder^146^, drinking^145^, smoking^145^, cannabis use^147^, general happiness^148^, snoring^149^, insomnia^149^, alcohol dependence^150^). PRS-CSx, a high-dimensional Bayesian regression framework that places continuous shrinkage prior on single nucleotide polymorphisms effect sizes^151^, was applied to enhance cross-population prediction. This method has consistently shown superior performance compared to other methods across a wide range of genetic architectures in simulation and real data analyses^151^. Hyperparameter optimization for the GPSs was conducted using a held-out validation set of 1,579 unrelated participants. Adjustments for population stratification were performed based on the first ten ancestrally informative principal components to account for potential confounding effects.

#### Anatomical Brain Imaging: T1/T2, Freesurfer 6

Baseline year T1-weighted (T1w) 3D structural MRI acquired in the ABCD study were processed following established protocols^152,153^: To maximize geometric accuracy and image intensity reproducibility, gradient nonlinearity distortion was corrected^154^. After correcting intensity nonuniformity using tissue segmentation and spatial smoothing, images were resampled to 1 mm isotropic voxels. We used Freesurfer v6.0 (https://surfer.nmr.mgh.harvard.edu) for the following procedures: cortical surface followed by skull-stripping^155^, white matter segmentation, and mesh creation^156^, correction of topological defects, surface optimization^157^, and nonlinear registration to a spherical surface-based atlas^158,159^. Using Desikan–Killiany atlas^160^, a standard atlas for Freesurfer and ABCD study, we extracted 399 brain ROI measures, including volumes, surface area, thickness, mean curvature, sulcal depth, and gyrification.

#### Functional MRI (fMRI): Monetary Incentive Delay (MID) task

The MID task was used measure the neural activation during anticipation and receipt of monetary gains and losses. In each trial, participants were shown a graphical cue of the 5 possible incentive types: large reward ($5), small reward ($0.20), large loss (-$5), small loss (-$0.20), or neutral ($0). The incentive cue is presented for 2,000 ms, followed by a jittered anticipatory delay (1,500–4,000 ms). Subsequently, a target to which participants respond to gain or avoid losing money was shown (150–500 ms), and feedback of their performance was provided (2,000 ms). A total of 40 reward, 40 loss, and 20 neutral trials were presented in pseudo-random order across the two task runs. Task parameters was dynamically manipulated for each subject to maintain 60% success rate^152^. We used baseline year observations of average beta weights of the MID task fMRI with Desikan-Killiany parcellations^160^.

#### Covariates

To adjust for the potential confounding effects, sociodemographic covariates were included. Consistent with existing research on psychiatric disorders in ABCD samples^3,114,128,161^, we controlled for the child’s sex, age, race/ethnicity, caregiver’s relationship to a child, BMI, parental education, marital status of the caregiver, household income, parent’s age, and family history of psychiatric disorders. The family history of psychiatric disorders, measured as the proportion of first-degree relatives who experienced psychosis, depression, mania, suicidality, previous hospitalization, or professional help for mental health issues^3^ was included as a covariate. Given that delay discounting and PLEs are associated with an individual’s neurocognitive capabilities^162–164^, NIH Toolbox total intelligence was used as a covariate. All covariates were from baseline year observations.

### Statistical Analyses

#### Instrumental Variable Regression

The IV method controls unobserved confounding bias by identifying an instrumental variable Z which causally affects the independent variable of interest X but has no direct effect on the dependent variable Y^60^. Our instrument variable for ADI was a variable indicating whether the state in each subject resides at baseline year has legislation prohibiting discrimination by the source of income (SOI laws) in the housing market. According to a report by the US Department of Housing and Urban Development, landlords accept housing vouchers 20.2%p∼59.3%p higher in local areas with SOI laws^165^. Research shows significant reductions in neighborhood poverty rates in locations with SOI laws^166^, and those who receive the benefits of housing vouchers in childhood show lower hospitalization rates, less impulsive consumption^167^, and substantially better mental health^168^. Taken together, we hypothesized that living in states with SOI laws would lead to more moderate discounting of future rewards and fewer PLEs, only through a positive influence on the neighborhood socioeconomic conditions of the subjects.

F-statistic above ten is considered a strong instruments^169^. The F-statistic for each model was F= 34.031 (p<0.0001), suggesting that our IV model is not likely to suffer from weak instrument bias. Also, testing endogeneity of ADI (i.e., whether ADI as a treatment variable or predictor correlates with the error term), we found that the model was significantly biased by unobserved confounding (all Hausman test^170^ for differences, p≤0.0158). This justifies the need for the IV regression approach to control for the significant confounding effects and to test the causal relationship of neighborhood disadvantage with delay discounting and psychopathology. All continuous variables were standardized (z-scaled), and analyses were run using *ivreg*^171^ in R version 4.1.2. For all analyses, threshold for statistical significance was set at p<0.05, with multiple comparison correction based on false discovery rate.

#### Causal Machine Learning for Treatment Effects

IV Forest (*grf* R package version 2.2.1)^57,58^ is a novel causal machine learning approach extends from the conventional random forest framework^172^ with recursive partitioning, subsampling, and random splitting to identify the average treatment effects and its individual differences. We obtained augmented inverse propensity weighted estimates of average treatment effects, a doubly-robust estimator which can capture complex patterns of individual differences and do not rely on a priori model assumptions^57^ such as linearity. This is particularly advantageous when the relationship between environmental variables and neurocognitive development is likely nonlinear^7,173,174^. To measure the average outcome between treated versus untreated subjects, ADI was binarized (i.e., mean split).

In line with prior studies^67,68^, we evaluated heterogeneous treatment effects by testing whether the average treatment effects are significantly different among subgroups defined by their relative resilience/vulnerability^69^. These subgroups were defined across a decile spectrum, with Q1 representing the most vulnerable and Q10 the most resilient. We considered a model to have significant heterogeneous treatment effects only if it satisfied all three of the following criteria:

i. The monotonicity test evaluates the existence of at least one inequality in the average treatment effects across the deciles. This is achieved by whether to accept or reject the null hypothesis (J-f0), which states that treatment effects are equal across all deciles. Essentially, the test determines whether there is a consistent, ordered relationship in the treatment effects from one decile to the next, indicating a monotonic trend.
ii. The alternative hypothesis test evaluates whether the average treatment effect in the highest decile exceeds the combined average treatment effects in the remaining deciles Q2 through Q10.
iii. The ANOVA test determines whether the average treatment effects are statistically different across deciles. In this context, the group mean in the ANOVA corresponds to the average treatment effect of each decile.

To ensure that the IV Forest estimations are robust across different random seeds, we developed 100-seed ensemble IV Forest model. Specifically, we used the following procedures:

1. For each iteration, randomly split the data in half (i.e., train vs test sets) to build a forest model with the first half and perform estimation with the other half. We repeated this process 100 times using different seeds in each iteration to build 100 forest models.
2. Combine the 100 forest models into one big IV Forest model and then rank the observations into deciles according to their estimated conditional average treatment effects.
3. Obtain augmented inverse propensity weighted average treatment effects for each decile and perform monotonicity, alternative hypothesis, and ANOVA tests.

## Data Availability

All data produced in the present work are contained in the manuscript

## Acknowledgements

This work was supported by the National Research Foundation of Korea (NRF) grant funded by the Korea government (MSIT) (No. 2021R1C1C1006503, 2021K1A3A1A2103751212, 2021M3E5D2A01022515, RS-2023-00266787, RS-2023-00265406), by Creative-Pioneering Researchers Program through Seoul National University (No. 200-20230058), by Semi-Supervised Learning Research Grant by SAMSUNG (No.A0426-20220118), by Identify the network of brain preparation steps for concentration Research Grant by LooxidLabs (No.339-20230001), and by Institute of Information & Communications Technology Planning & Evaluation (IITP) grant funded by the Korea government (MSIT) [NO.2021-0-01343, Artificial Intelligence Graduate School Program (Seoul National University)]

## Supplementary Information

**Supplementary Fig. 1.**
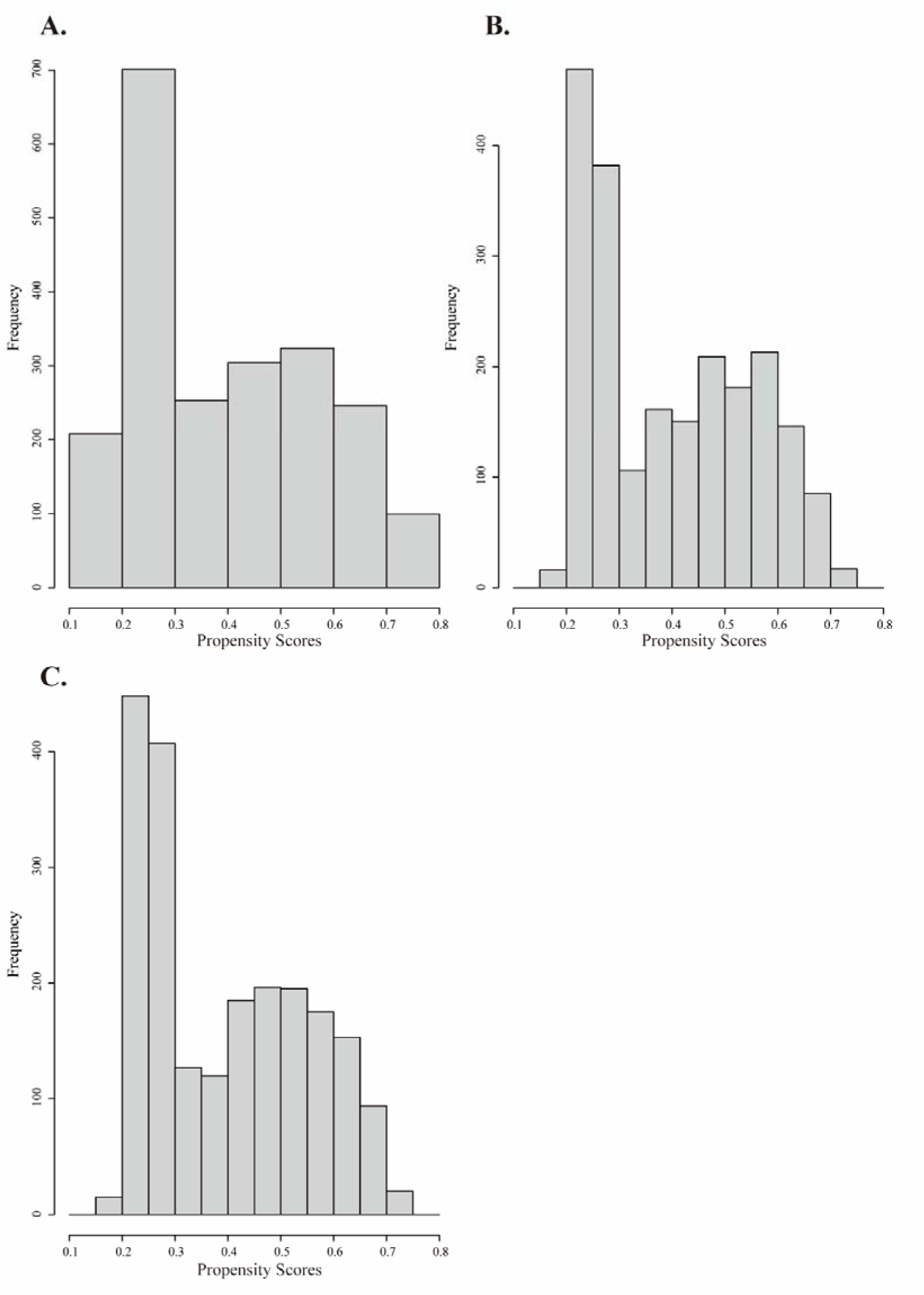
Assessment of the overlap assumption. Histograms of propensity scores for the three distinct IV Forest models: Delay Discounting model (**A**), Gene-Brain model (**B**), and Integrated model (**C**). The IV Forest algorithm relies on the overlap assumption, which posits that there should be sufficient overlap in the covariate distributions between the treated and control groups. In other words, this means that the treatment status of an individual should not be deterministically predictable based on their covariates. The validity of the overlap assumption is evaluated using the histograms of propensity scores^1,2^. The fact that none of the estimated propensity scores are extremely close to either one or zero indicates that the overlap assumption holds, suggesting an appropriate level of randomness in treatment assignment relative to the covariates.

**Supplementary Table 1.** Comparison of the sample and national demographics. Since Household income for subjects in the study is presented as deciles, it is transformed into a monetary value by considering the income limits for each decile. The data for the US national demographics is available at Data is available at https://www.census.gov/en.html.

|  |  | Mean or Ratio (%) |  |
| --- | --- | --- | --- |
|  |  | Subjects in our study | US National Demographics |
| Family Income | | \$70,245 | \$60,336 |
| Sex | Male | 53.86% | 51.16% |
| Ethnicity/Race | White | 65.57% | 57.8% |
|  | Black | 6.37% | 12.1% |
|  | Hispanic | 17.47% | 18.7% |
|  | Asian | 0.33% | 6.1% |
|  | Other | 10.26% | 12.4% |

**Supplementary Table 2.** Results of linear IV regression. Conventional linear IV regression was conducted to confirm the average treatment effects of neighborhood socioeconomic adversity on children’s delay discounting and psychotic risk. All p-values were corrected for multiple comparison using false discovery rate.

|  |  | Conventional Linear IV Regression |  |  |  |  |
| --- | --- | --- | --- | --- | --- | --- |
|  |  | Estimates | Std. Error | 95% Lower CI | 95% Upper CI | P-FDR |
| Delay Discounting |  | -0.468 | 0.216 | -0.891 | -0.045 | 0.03 |
| PLEs | Distress Score PLE (1-year follow-up) | 0.609 | 0.224 | 0.170 | 1.047 | 0.011 |
|  | Distress Score PLE (2-year follow-up) | 0.780 | 0.234 | 0.320 | 1.239 | 0.003 |
|  | Delusional Score PLEs (1-year follow-up) | 0.486 | 0.215 | 0.066 | 0.906 | 0.028 |
|  | Delusional Score PLEs (2-year follow-up) | 0.578 | 0.223 | 0.141 | 1.015 | 0.013 |
|  | Hallucinational Score PLEs (1-year follow-up) | 0.604 | 0.221 | 0.172 | 1.036 | 0.011 |
|  | Hallucinational Score PLEs (2-year follow-up) | 0.827 | 0.241 | 0.354 | 1.299 | 0.003 |

### Double ML models

To confirm robustness of the IV Forest results, we used double machine learning (Double ML). This up-to-date causal machine learning method can utilize any state-of-the-art machine learning models to obtain consistent, unbiased estimates of average treatment effects by partialling out the confounding effects of covariates^3^. It is particularly effective when the covariates are high-dimensional and have complex interactions.

We used partial linear model instrumental variable model. In the partial linear model, we only assume linearity of the treatment variable ADI while the relationships between the outcome variable Y and covariates X and between instrument variable SOI and covariates X remain as an unknown function. Below shows the simple mathematical representation of the model:

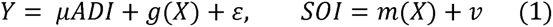

Here, Y denote for outcome variable (in our study, delay discounting and psychotic-like experiences), ADI the treatment variable, X multidimensional covariates, SOI the instrument variable. g(), m() are unknown functions and E, v are random errors. In the partial linear model (Equation 1), we assume that the treatment variable (i.e., ADI) have linear relationship with the outcome variable Y. There are no model assumptions specifying the relationship between multidimensional covariates X, outcome Y, and the instrumental variable SOI.

We built an ensemble machine learning pipeline consisting of elastic net, random forest, XGBoost, and support vector machine with parameters tuned via 10-fold cross validation. In general, ensemble methods can improve model performance with lower error and higher accuracy by combining several base models^4^. For each analysis, all continuous variables were standardized (z-scaled) beforehand to obtain standardized estimates, and analyses were run using *DoubleML* packages^5^ in R version 4.1.2.

**Supplementary Table 3.** Results of Double ML models. All p-values were corrected for multiple comparison using false discovery rate.

|  | **Double ML Partial Linear IV Regression** |  |  |  |  |
| --- | --- | --- | --- | --- | --- |
|  | **Estimates** | **Std. Error** | **95% Lower CI** | **95% Upper CI** | **P-FDR** |
| Delay Discounting<br>(1-year follow-up) | -0.429 | 0.206 | -0.834 | -0.025 | 0.044 |
| Distress Score PLE<br>(1-year follow-up) | 0.495 | 0.199 | 0.105 | 0.884 | 0.023 |
| Distress Score PLE<br>(2-year follow-up) | 0.609 | 0.193 | 0.232 | 0.987 | 0.005 |
| Delusional Score PLEs<br>(1-year follow-up) | 0.393 | 0.201 | -0.001 | 0.788 | 0.051 |
| Delusional Score PLEs<br>(2-year follow-up) | 0.417 | 0.196 | 0.032 | 0.801 | 0.044 |
| Hallucinational Score PLEs<br>(1-year follow-up) | 0.498 | 0.187 | 0.131 | 0.864 | 0.018 |
| Hallucinational Score PLEs<br>(2-year follow-up) | 0.683 | 0.188 | 0.316 | 1.051 | 0.002 |

### Random Forest-based Feature Selection using Boruta algorithm

We used Boruta^6^ to select GPS and brain ROIs of structural MRI and MID task fMRI significantly associated with delay discounting. Boruta first generates shadow attributes, which is irrelevant to the outcome, by shuffling all input features. It then confirms features that have significantly higher importance in predicting the outcome than the shadow attributes with 95% confidence level, Bonferroni-corrected two-tailed tests^6^. The selected features were included as covariates in the IV Forest models for assessing heterogeneous treatment effects of ADI.

**Supplementary Table 4.**
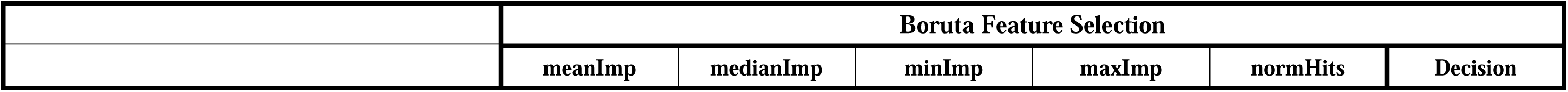
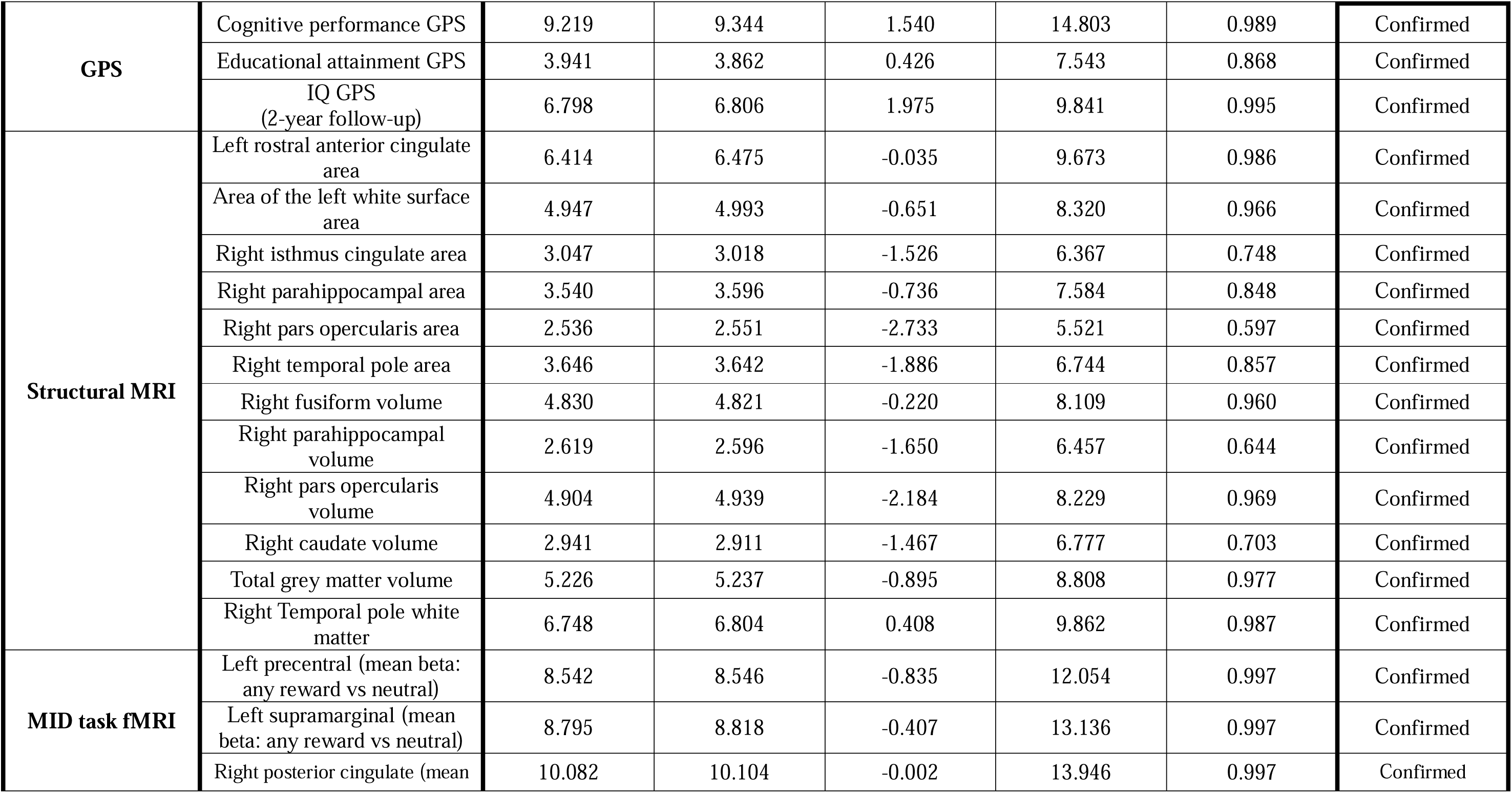
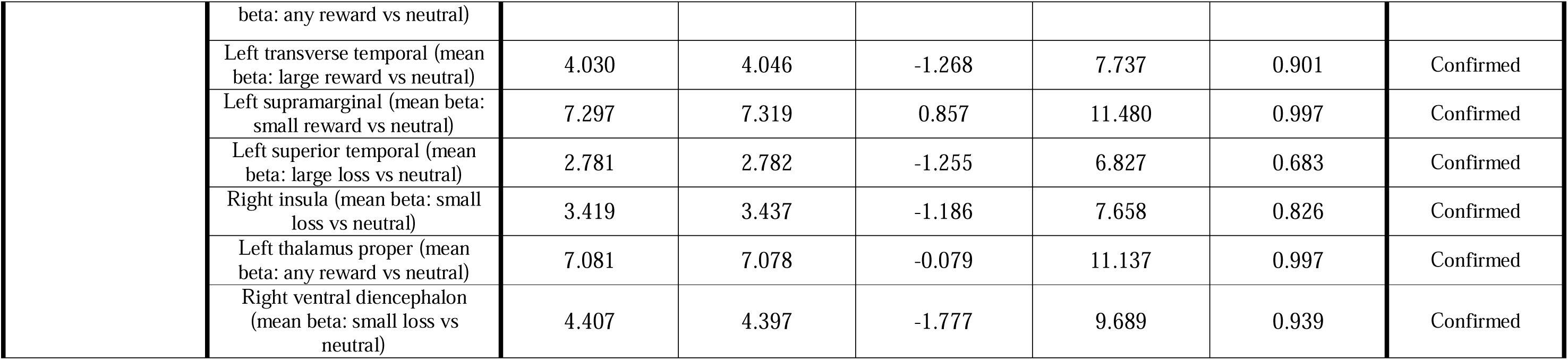
Genetic and neural correlates of delay discounting selected using random forest-based algorithm. Using the Boruta algorithm, we selected polygenic scores, neuroanatomical, and MID task functional activations related to delay discounting. meanImp, medianImp, minImp, and maxImp denote mean, median, maximal, and minimal importance, respectively. normHits indicate the number of hits normalized to number of importance source runs performed. *Decision* indicates the final decision of whether or not to select each feature.

|  | Boruta Feature Selection |  |  |  |  |  |
| --- | --- | --- | --- | --- | --- | --- |
|  | meanImp | medianImp | minImp | maxImp | normHits | Decision |

|  |  |  |  |  |  |  |  |
| --- | --- | --- | --- | --- | --- | --- | --- |
| **GPS** | Cognitive performance GPS | 9.219 | 9.344 | 1.540 | 14.803 | 0.989 | Confirmed |
|  | Educational attainment GPS | 3.941 | 3.862 | 0.426 | 7.543 | 0.868 | Confirmed |
|  | IQ GPS<br>(2-year follow-up) | 6.798 | 6.806 | 1.975 | 9.841 | 0.995 | Confirmed |
| **Structural MRI** | Left rostral anterior cingulate area | 6.414 | 6.475 | -0.035 | 9.673 | 0.986 | Confirmed |
|  | Area of the left white surface area | 4.947 | 4.993 | -0.651 | 8.320 | 0.966 | Confirmed |
|  | Right isthmus cingulate area | 3.047 | 3.018 | -1.526 | 6.367 | 0.748 | Confirmed |
|  | Right parahippocampal area | 3.540 | 3.596 | -0.736 | 7.584 | 0.848 | Confirmed |
|  | Right pars opercularis area | 2.536 | 2.551 | -2.733 | 5.521 | 0.597 | Confirmed |
|  | Right temporal pole area | 3.646 | 3.642 | -1.886 | 6.744 | 0.857 | Confirmed |
|  | Right fusiform volume | 4.830 | 4.821 | -0.220 | 8.109 | 0.960 | Confirmed |
|  | Right parahippocampal volume | 2.619 | 2.596 | -1.650 | 6.457 | 0.644 | Confirmed |
|  | Right pars opercularis volume | 4.904 | 4.939 | -2.184 | 8.229 | 0.969 | Confirmed |
|  | Right caudate volume | 2.941 | 2.911 | -1.467 | 6.777 | 0.703 | Confirmed |
|  | Total grey matter volume | 5.226 | 5.237 | -0.895 | 8.808 | 0.977 | Confirmed |
|  | Right Temporal pole white matter | 6.748 | 6.804 | 0.408 | 9.862 | 0.987 | Confirmed |
| **MID task fMRI** | Left precentral (mean beta: any reward vs neutral) | 8.542 | 8.546 | -0.835 | 12.054 | 0.997 | Confirmed |
|  | Left supramarginal (mean beta: any reward vs neutral) | 8.795 | 8.818 | -0.407 | 13.136 | 0.997 | Confirmed |
|  | Right posterior cingulate (mean beta: any reward vs neutral) | 10.082 | 10.104 | -0.002 | 13.946 | 0.997 | Confirmed |
|  | Left transverse temporal (mean beta: large reward vs neutral) | 4.030 | 4.046 | -1.268 | 7.737 | 0.901 | Confirmed |
|  | Left supramarginal (mean beta: small reward vs neutral) | 7.297 | 7.319 | 0.857 | 11.480 | 0.997 | Confirmed |
|  | Left superior temporal (mean beta: large loss vs neutral) | 2.781 | 2.782 | -1.255 | 6.827 | 0.683 | Confirmed |
|  | Right insula (mean beta: small loss vs neutral) | 3.419 | 3.437 | -1.186 | 7.658 | 0.826 | Confirmed |
|  | Left thalamus proper (mean beta: any reward vs neutral) | 7.081 | 7.078 | -0.079 | 11.137 | 0.997 | Confirmed |
|  | Right ventral diencephalon (mean beta: small loss vs neutral) | 4.407 | 4.397 | -1.777 | 9.689 | 0.939 | Confirmed |

### Mediation Analysis

To test the role of delay discounting as a mediator between ADI and PLEs, we also used a linear mediation analysis. By utilizing *mediation* package^7^ in R, we conducted causal mediation analysis by decomposing local average treatment effect (LATE) into local average causal mediation effect (LACME) and local average natural direct effect (LANDE). In order to control unobserved confounding bias, *ivmediate* function was utilized to incorporate the instrument variable in the causal mediation analysis. In this analysis, ADI and delay discounting were transformed as a binary variable (i.e., above or below mean).

**Supplementary Table 5.**
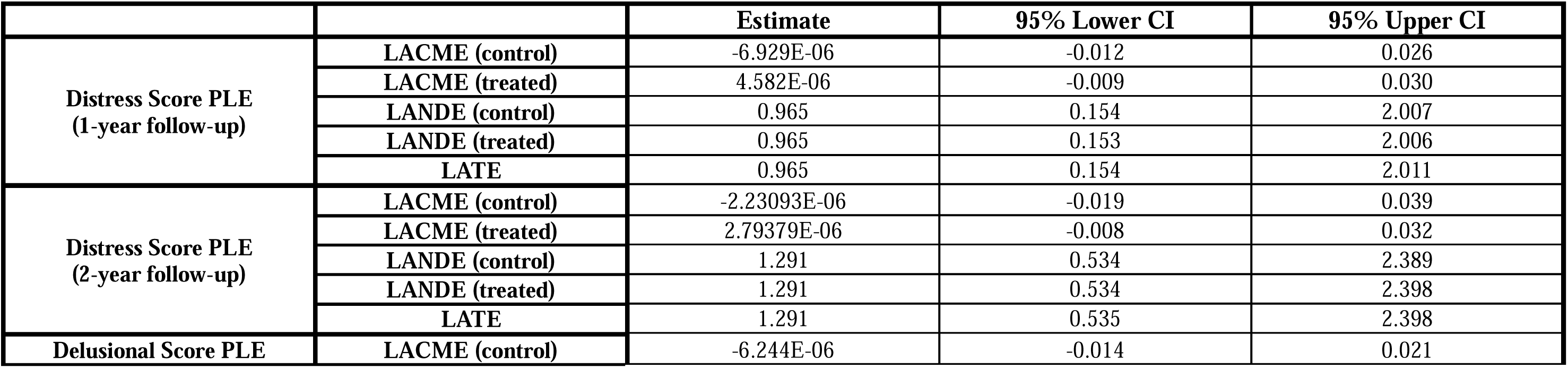
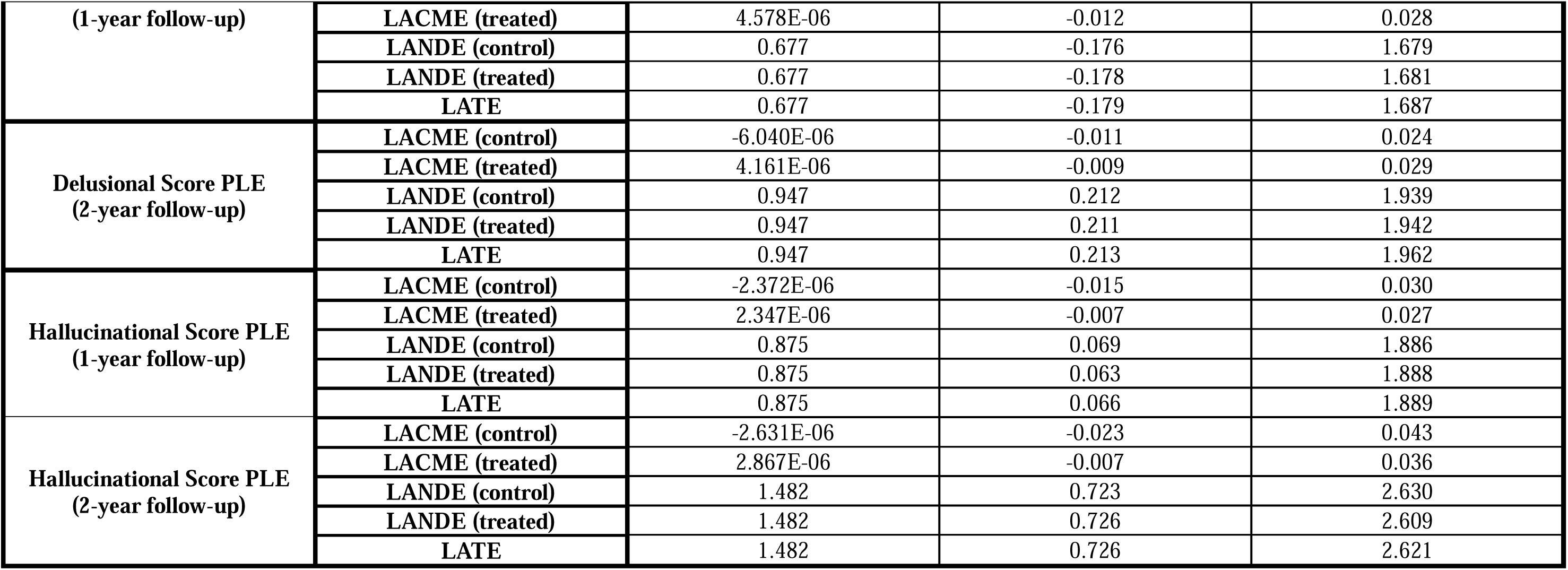
Results of conventional linear IV mediation. LACME represents the average hypothetical change in the outcome among compliers when the mediator is changed from the value under the treatment status to the control status while the treatment variable is fixed. LANDE represents the average hypothetical change in the outcome among compliers when the treatment variable is changed from the treatment status to the control status while the mediator is fixed.

